# Telomerase Reverse Transcriptase Enzyme (hTERT) expression in bronchial mucosa inversely correlates with the severity of airway obstruction in patients with chronic airway obstructive diseases

**DOI:** 10.1101/2022.12.08.22283249

**Authors:** C. Moermans, L. Medard, S. Graff, MS. Njock, N. Bougard, F. Schleich, J. Guiot, J-L. Corhay, R. Louis

**Affiliations:** Giga I3 research group, Liege University, Liege, Belgium; Dept. of Pneumology-Allergology, CHU of Liege, 4000 Liege, Belgium

**Keywords:** Asthma, COPD, bronchial biopsies, Telomerase, hTERT

## Abstract

**Rationale:** Asthma and Chronic Obstructive Pulmonary Disease (COPD) are the two main chronic airway inflammatory diseases related to aging. Cellular aging is prevented by telomeres and their shortening induces cellular senescence. Human Telomerase Reverse Transcriptase Enzyme (hTERT) can act directly on DNA and prevent telomere shortening. The goal of this study was to assess hTERT expression in the bronchial mucosa of patients suffering from chronic airway obstructive diseases and to relate the hTERT expression to demographics and lung function characteristics.

**Methods:** We collected bronchial biopsies from 38 patients suffering from chronic airway diseases including 21 severe asthmatics and 17 COPD who underwent bronchoscopy in routine practice. hTERT expression was assessed by immunochemistry and co-immunostaining was performed to identify hTERT positive cells within the different immune cell populations.

**Results:** hTERT expression in airway mucosa was essentially found in structural cells. Among leukocytes, lymphocytes were the principal cells expressing hTERT. On the whole cohort, hTERT expression score was not different between men and women and not influenced by the age nor by the smoking history as reflected by the pack years. Total airway hTERT positive staining score positively correlated with post-bronchodilation (post-BD) FEV_1_ % predicted and FEV_1_/FVC. (r=0.38, p<0.05 for both). hTERT+ lymphocytes were also positively correlated to post-BD FEV_1_/FVC (r=0.37, p<0.05). Consequently, hTERT expression was lower in those patients with fixed airway obstruction (post-BD FEV1/FVC < 70%). There was no difference between asthmatics and COPD.

**Conclusions:** In patients with chronic airway diseases, total and lymphocyte hTERT expressions inversely correlate with the degree of airway obstruction and are reduced in patients with fixed airway obstruction, which supports a role of hTERT in airway remodeling.

## INTRODUCTION

As life expectancy is increased globally, the number of people over the age of 65 years is estimated to nearly triple (1.5 billion) by 2050 representing 16% of the world’s population[1]. This aging population is living longer with chronic diseases, including asthma and Chronic Obstructive Pulmonary Disease (COPD), the two main chronic airway diseases worldwide. Chronic lower respiratory diseases are also the third leading cause of death for elderly people[2].

One of the defining mechanisms behind cellular aging is related to telomere shortening[3]. Telomeres are repetitive sequences TTAGGG in humans[4] which decrease at each cell cycle[5] to induce progressively cellular apoptosis[6].

To avoid telomere shortening, the telomerase can act directly on DNA and add new telomeres at the end of chromosomes. Telomerase contains 2 main subunits: a telomerase RNA component (TERC) and the catalytic subunit of telomerase reverse transcriptase (TERT) [7]. Particularly for asthma and COPD, several studies showed that the circulating leukocytes have reduced telomere lengths in correlation with disease severity and lung function [8,9]. Although, pulmonary vascular endothelial cells as well as alveolar epithelial cells were reported to have shortened telomeres [10] in COPD, literature on telomere length or telomerase expression in airways, the primarily affected area of asthma and COPD, appeared limited. To our knowledge, only one study reported decreased TERT on bronchial biopsies in patients with asthma and COPD compared to controls [8]. However, the affected leukocyte type was not highlighted neither a potential link with airway obstruction.

Our study aimed to analyze the TERT expression in bronchial biopsies of patients with airway obstructive diseases by combining asthma and COPD and to investigate the relationship between TERT immunostaining and patient’s demographic and functional characteristics.

## MATERIALS AND METHOD

### 1. Patients

Thirty-eight patients were recruited from the CHU of Liege, Belgium. This study took place from September 2017 to March 2020 and all patients gave consent to participate. They were all patients who had been admitted for bronchoscopy performed for diagnosis purpose. Twenty-one asthmatics and 17 COPD patients were included in this study. Asthma was diagnosed according to the Global Initiative for Asthma guidelines (http://ginasthma.org/). Bronchoscopy in asthma context was processed in case of incoercible cough, tumor suspicion or thoracic image abnormalities. COPD diagnosis was made according to the GOLD criteria (http://goldcopd.org/). For all patients, fiberoptic bronchoscopy was performed under local anesthesia and mild sedation through IV midazolam. After visual inspection of the airways, 3-4 biopsies were taken from a lobar bronchus not suspected of tumor infiltration when the bronchoscopy was indicated for tumor suspicion. The specimens came from residual material. This study was approved by the Ethics committee of CHU Liege (2015/00c).

### 2. Respiratory function

Fractional Exhaled Nitric Oxide (FeNO) was measured using NiOX (Aerocrine, Solna, Sweden) at a flow rate of 50 mL/s. Spirometry was performed (forced expiratory volume in 1 second [FEV_1_] and forced vital capacity [FVC] maneuver) according to the American Thoracic Society (ATS)/European Respiratory Society (ERS) standard criteria [11]. Blood samples of patients were analyzed by the routine laboratory of the University Hospital of Liege.

### 3. Immunohistochemistry (IHC) and image analysis

The specimens were fixed in 4% formalin and routinely processed to paraffin blocks. These specimens were treated by our immune-histology platform. Specimens were cut into 5 μm slices by a microtome. Immunohistochemistry was performed on paraffin-embedded bronchial biopsies to evaluate expression of human TERT (hTERT). After antigen unmasking of the slices by citrate and heating, and blocking unspecific background with Protein Block Serum Free (X090930-2, Dako, CA, USA), incubation with the primary and secondary antibodies was performed. We used specific antibodies against hTERT (1:200, ab230527, abcam, Cambridge, UK), neutrophils (ELA 1:2000, MAB 91671-100, R&D systems, Minneapolis, USA), lymphocytes (CD3, 1:250, RM9107-S, Fisher Scientific, Hampton, USA) and macrophages (CD68, 1:20, 790-2931, Roche, Penzberg, Germany). The color reaction was revealed using Liquid DAB and Substrate Chromogen System (K3468, Dako, CA, USA) and High Def Red IHC (ADI-950-I41-0030, Enzo, NY, USA). Subsequently, hematoxylin staining was carried out. The identification of eosinophils was performed morphologically after hematoxylin eosin staining (see online supplementary material). Structural cells represented a combination of fibrocytes and vascular cells and were validated by a histopathologist.

The slides were then scanned on NDP NanoZoomer Digital Pathology scanner (Hamamatsu, Japan). The hTERT positive cells in the submucosa as well as and the immune cells were counted using QuPath open source digital software [12] and normalized by relative biopsy area.

### 4. Statistical analysis

Data were presented as mean ± SD, or median (interquartile range) for continuous variables according to the distribution of the data. Data from 2 independent groups were compared using an unpaired t test for normally distributed data and a Mann-Whitney test for continuous variables incompatible with a normal distribution. Correlations were performed with Spearman tests. A p value < 0.05 was considered statistically significant. GraphPad Prism 7 (Graphpad Software San Diego, CA, USA) was used for statistical analysis and graphic generation.

## RESULTS

Patient characteristics are displayed in Table 1.

**Table 1:** Demographic and functional characteristics.

| Characteristics | Obstructive airway diseases (n=38) | Asthma (n= 21) | COPD (n=17) |
| --- | --- | --- | --- |
| Sex (M/F) | 17/21 | 7/14 | 10/7 |
| Age (years) | 61 ± 11 | 57 ± 10 | 66 ± 10** |
| BMI (kg/m<sup>2</sup>) | 25 ± 4 | 26 ± 4 | 24 ± 5 |
| Tobacco status<br>NS/CS/EXS | 6/12/20 | 6/2/13 | 0/10/7** |
| Pack Year | 26 (10-40) | 10 (0-27) | 39 (29-60)*** |
| FEV<sub>1</sub> pre BD (% predicted) | 62 ± 21 | 68 ± 23 | 55 ± 17 |
| FEV<sub>1</sub>/FVC pre BD (%) | 65 ± 13 | 70 ± 13 | 59 ± 10** |
| TLC (% predicted) | 95 ± 15 | 92 ± 14 | 97 ± 16 |
| RV (% predicted) | 139 ± 40 | 135 ± 44 | 145 ± 35 |
| DLCO (% predicted) | 58 ± 16 | 64 ± 16 | 53 ± 15 |
| KCO (% predicted) | 77 ± 20 | 87 ± 19 | 68 ± 15** |
| Blood leukocyte count (10<sup>3</sup>/μL) | 8.4 (7.1-10.8) | 8.5 (7.1-10.5) | 8.2 (6.9-11.9) |
| Blood eosinophil count (/μL) | 87 (42-201) | 85 (50-242) | 87 (26-201) |
| Blood neutrophil count (/μL) | 5345 (4263-7384) | 4648 (3913-7565) | 5897 (4848-7361) |
| ICS (%) | 23 (61) | 18 (86) | 5 (29)*** |
| OCS (%) | 6 (16) | 4 (19) | 2 (12) |
| LABA (%) | 25 (66) | 18 (86) | 7 (41)** |
| LAMA (%) | 12 (32) | 5 (24) | 7 (41) |
| SABA (%) | 6 (16) | 5 (24) | 1 (6) |
| SAMA (%) | 14 (37) | 9 (43) | 5 (29) |
| LTRA (%) | 9 (24) | 7 (33) | 2 (12) |
| Biotherapy |  |  |  |
| Anti Il-5 (%) | 3 (8) | 3 (14) | 0 (0) |
| Anti IgE (%) | 2 (5) | 2 (10) | 0 (0) |

Results are presented as absolute number, absolute number (percentage), median (interquartile range) or mean ± SD; BMI: body mass index; NS: non-Smoker; CS: current smoker; ExS: ex- smoker; FEV<sub>1</sub>: forced expiration volume in 1s; BD: bronchodilation; TLC: total lung capacity; RV: residual volume; DLCO: carbon monoxide diffusing capacity; KCO: carbon monoxide transfer coefficient; ICS: inhaled corticosteroids; OCS: oral corticosteroids, LABA: long acting beta 2 agonist, LAMA: long acting muscarinic antagonist. SABA: short acting beta agonist; SAMA: short acting muscarinic antagonist; LTRA: leukotriene receptor antagonist. \*: P-value < 0.05; \*\*: P-Value < 0.01; \*\*\* P-Value < 0.001 vs asthmatics.

### a) hTERT expression within bronchial mucosa

HTERT staining results are detailed in Table 2. In both asthmatics and COPD structural cells (non-leukocytes) were the majority of cells identified in the bronchial mucosa and 3% of these structural cells were hTERT positive. Among leukocytes, lymphocytes represented the majority of immune cells identified in bronchial mucosa and 11 to 15% of lymphocytes stained for hTERT. Neutrophils were the second most frequent leukocyte type observed within bronchial biopsies but to a much lower extent than lymphocytes and only 0-3% of neutrophils stained for hTERT. Quantity of macrophages and eosinophils were negligible. Overall there was no difference in the hTERT staining between asthmatics and COPD.

**Table 2:** HTERT staining results in Asthma (n=21) and COPD (n=17) patients.

|  | **Asthma** | **COPD** |
| --- | --- | --- |
| Total cell number/ mm<sup>2</sup> | 1789 (1081-2211) | 1376 (845-1920) |
| Total hTERT positive cell number (hTERT+/mm<sup>2</sup>) | 74 (42-111) | 66 (28-130) |
| Structural cells (number/ mm<sup>2</sup>) | 1456 (978-1775) | 1077 (787-1362) |
| hTERT positive structural cell number /mm<sup>2</sup> | 34 (12-78) | 24 (13-68) |
| % of hTERT positive structural cells<br>(structural cells hTERT+)/ structural cells | 3 (1-6) | 3 (1-8) |
| Lymphocyte number (CD3+/mm<sup>2</sup>) | 221 (107-456) | 212 (92-333) |
| hTERT positive lymphocyte number (CD3+hTERT+/mm<sup>2</sup>) | 34 (11-52) | 23 (11-63) |
| % of hTERT positive lymphocytes (CD3+ hTERT+)/CD3+ | 15 (7-19) | 11 (7-34) |
| Neutrophil number (ELA+/mm<sup>2</sup>) | 7 (2-15) | 8 (3-22) |
| hTERT positive neutrophil number (ELA+hTERT+/mm<sup>2</sup>) | 0.0 (0.0-0.4) | 0.1 (0.0-2.2) |
| % of hTERT positive neutrophils (ELA+ hTERT+)/ELA+ | 0 (0-4) | 3 (0-10) |
| Macrophage number (CD68+/mm<sup>2</sup>) | 0.0 (0.0-0.4) | 0.3 (0.0-3.2) |
| hTERT positive macrophage number (CD68+hTERT+/mm<sup>2</sup>) | 0.0 (0.0-0.0) | 0.0 (0.0-0.0) |
| % of hTERT positive macrophages<br>(CD68+ hTERT+)/CD68+ | 0 (0-0) | 0 (0-0) |
| Eosinophil number/mm<sup>2</sup> | 0.6 (0.0-2.0) | 1.9 (0.0-5.4) |
| hTERT positive eosinophil number /mm<sup>2</sup> | 0.0 (0.0-0.8) | 1.0 (0.0-2.0) |
| % of hTERT positive eosinophils<br>(eosinophil hTERT+)/eosinophil | 0 (0-45) | 33 (0-45) |

*Results (number of cells per mm<sup>2</sup> submucosa) are expressed as median (25%-75%).*

### b) Relationship with demographic characteristics

There was no difference in total and immune cell hTERT expressions between males and females and there was no significant correlation between age and total cell hTERT expression (r=0.27, p=0.10). Similarly, no correlation was observed between smoking pack years and hTERT staining, neither for total cells (r=-0.05, p=0.77) nor for lymphocytes (r=-0.004, p= 0.98).

### c) Relationship with lung function

We found a positive correlation between the total number of hTERT positive cells and post-bronchodilation FEV_1_ and FEV_1_/FVC ratio (r= 0.38, p< 0.05 for both). A similar correlation were found for hTERT positive lymphocytes number and pre- and post-bronchodilation FEV_1_/FVC ratios (r=0.34, p<0.05 and r=0.37, p< 0.05 respectively, Figure 1). In addition, we observed a significant correlation between the hTERT staining for the structural cell expressed as % and the FEV_1_ and CVF post-bronchodilation (r=0.36, p=0.05 and r=0.43, p<0.05 respectively).

**Figure 1:**
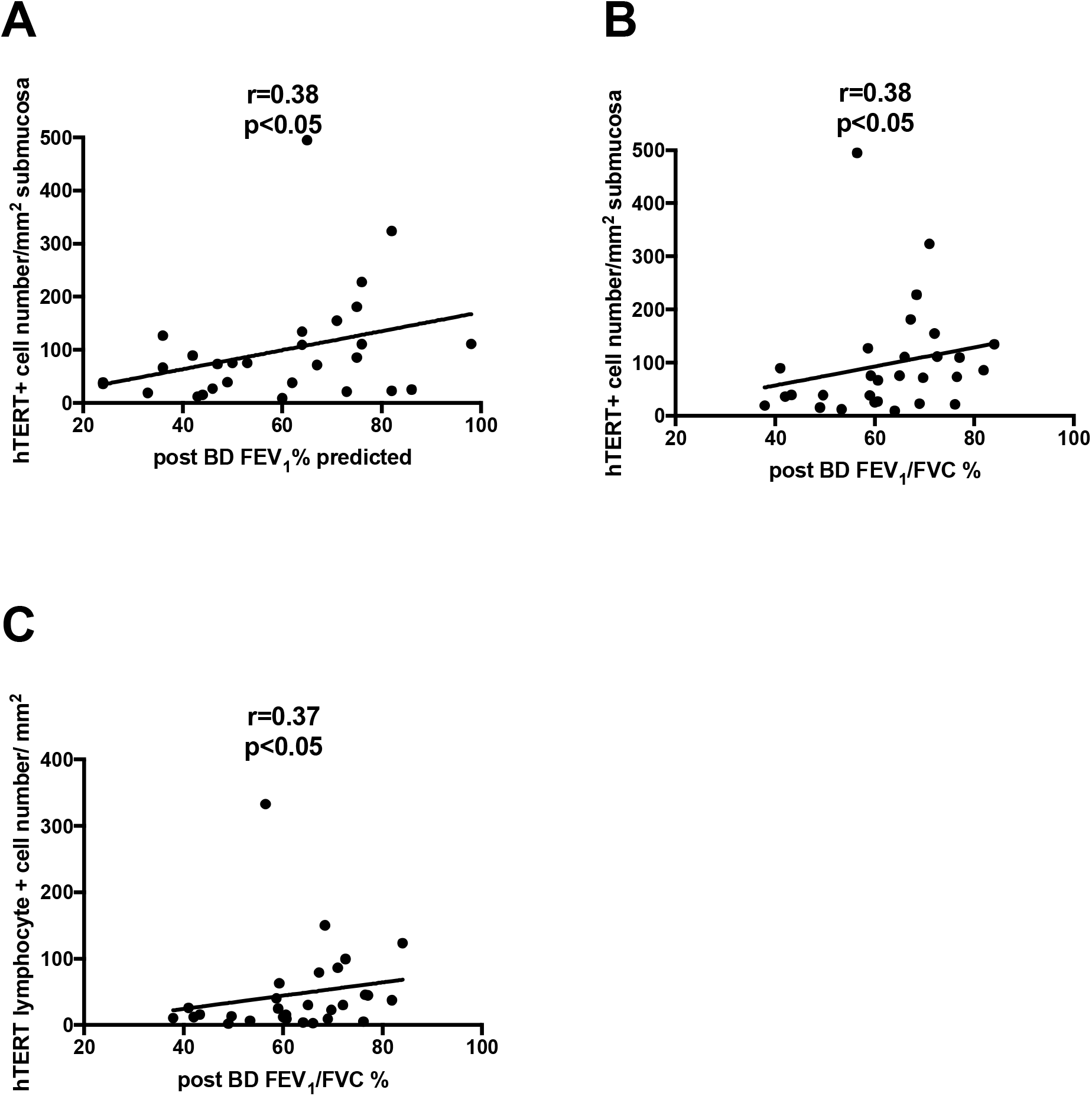
Correlation between hTERT protein expression in the submucosa of bronchial biopsies and lung function. **A** Correlation between the total number of hTERT positive cells and post-bronchodilation FEV1 expressed as predicted percentage. **B** Correlation between the total number of hTERT positive cells and post-bronchodilation FEV1/FVC. **C** Correlation between the hTERT positive lymphocyte number and post bronchodilation FEV1/FVC.

Among the 38 patients, 24 presented fixed airway obstruction defined by a post bronchodilation FEV_1_/FVC <70%. Total hTERT positive cell number as well as hTERT positive structural cell number were significantly decreased in biopsies of asthmatics and COPD patients with irreversible airway obstruction compared to others (p< 0.05 for both, Table 3 and Figure 2) with a trend for lower hTERT+ lymphocyte number (p=0.08).

**Table 3:**
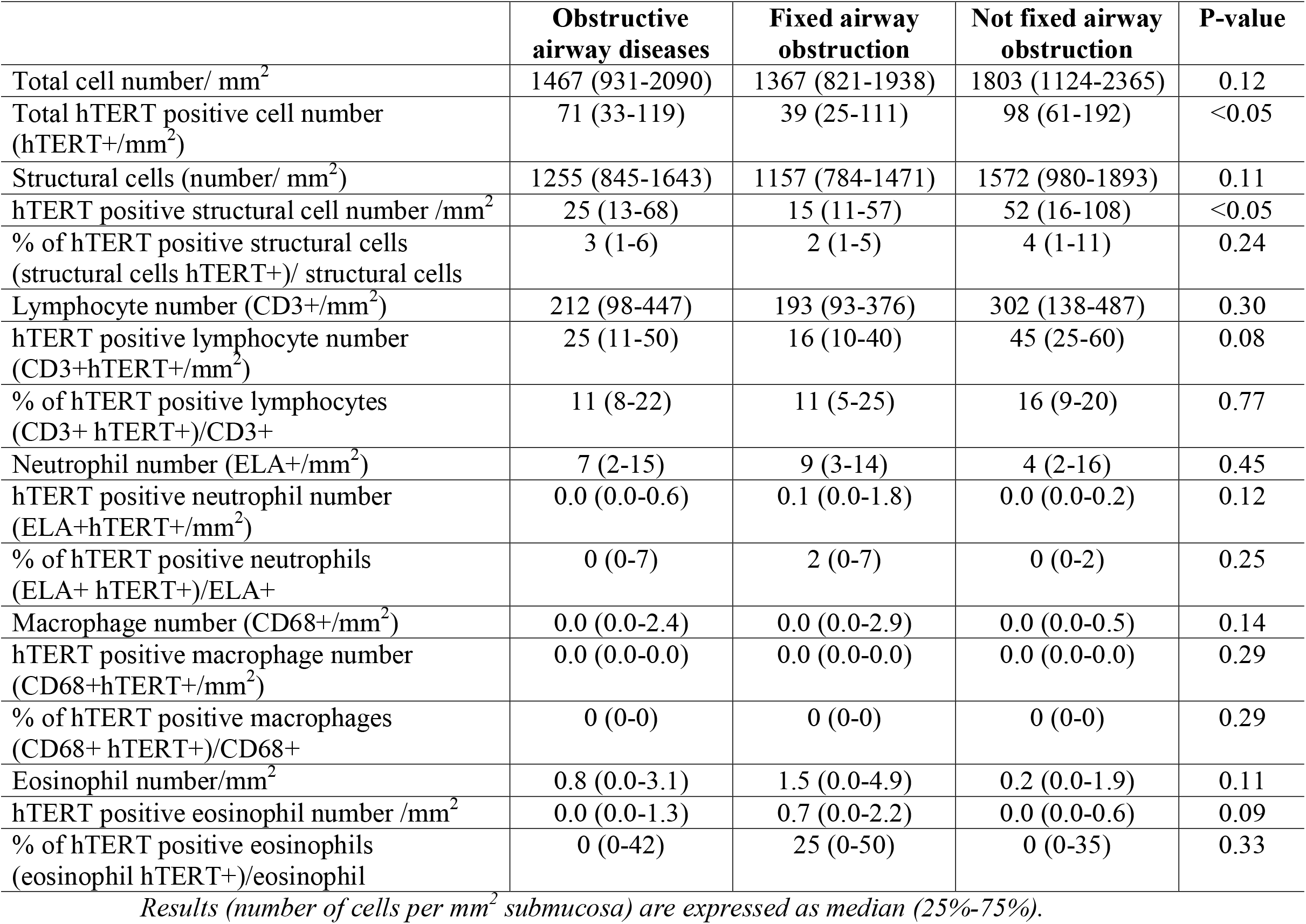
HTERT staining results in the whole cohort (n=38) and patients with fixed airway obstruction (n=24) vs not (n=14).

|  | **Obstructive**<br>**airway diseases** | **Fixed airway**<br>**obstruction** | **Not fixed airway**<br>**obstruction** | **P-value** |
| --- | --- | --- | --- | --- |
| Total cell number/ mm<sup>2</sup> | 1467 (931-2090) | 1367 (821-1938) | 1803 (1124-2365) | 0.12 |
| Total hTERT positive cell number<br>(hTERT+/mm<sup>2</sup>) | 71 (33-119) | 39 (25-111) | 98 (61-192) | <0.05 |
| Structural cells (number/ mm<sup>2</sup>) | 1255 (845-1643) | 1157 (784-1471) | 1572 (980-1893) | 0.11 |
| hTERT positive structural cell number /mm<sup>2</sup> | 25 (13-68) | 15 (11-57) | 52 (16-108) | <0.05 |
| % of hTERT positive structural cells<br>(structural cells hTERT+)/ structural cells | 3 (1-6) | 2 (1-5) | 4 (1-11) | 0.24 |
| Lymphocyte number (CD3+/mm<sup>2</sup>) | 212 (98-447) | 193 (93-376) | 302 (138-487) | 0.30 |
| hTERT positive lymphocyte number<br>(CD3+hTERT+/mm<sup>2</sup>) | 25 (11-50) | 16 (10-40) | 45 (25-60) | 0.08 |
| % of hTERT positive lymphocytes<br>(CD3+ hTERT+)/CD3+ | 11 (8-22) | 11 (5-25) | 16 (9-20) | 0.77 |
| Neutrophil number (ELA+/mm<sup>2</sup>) | 7 (2-15) | 9 (3-14) | 4 (2-16) | 0.45 |
| hTERT positive neutrophil number<br>(ELA+hTERT+/mm<sup>2</sup>) | 0.0 (0.0-0.6) | 0.1 (0.0-1.8) | 0.0 (0.0-0.2) | 0.12 |
| % of hTERT positive neutrophils<br>(ELA+ hTERT+)/ELA+ | 0 (0-7) | 2 (0-7) | 0 (0-2) | 0.25 |
| Macrophage number (CD68+/mm<sup>2</sup>) | 0.0 (0.0-2.4) | 0.0 (0.0-2.9) | 0.0 (0.0-0.5) | 0.14 |
| hTERT positive macrophage number<br>(CD68+hTERT+/mm<sup>2</sup>) | 0.0 (0.0-0.0) | 0.0 (0.0-0.0) | 0.0 (0.0-0.0) | 0.29 |
| % of hTERT positive macrophages<br>(CD68+ hTERT+)/CD68+ | 0 (0-0) | 0 (0-0) | 0 (0-0) | 0.29 |
| Eosinophil number/mm<sup>2</sup> | 0.8 (0.0-3.1) | 1.5 (0.0-4.9) | 0.2 (0.0-1.9) | 0.11 |
| hTERT positive eosinophil number /mm<sup>2</sup> | 0.0 (0.0-1.3) | 0.7 (0.0-2.2) | 0.0 (0.0-0.6) | 0.09 |
| % of hTERT positive eosinophils<br>(eosinophil hTERT+)/eosinophil | 0 (0-42) | 25 (0-50) | 0 (0-35) | 0.33 |

*Results (number of cells per mm<sup>2</sup> submucosa) are expressed as median (25%-75%).*

**Figure 2:**
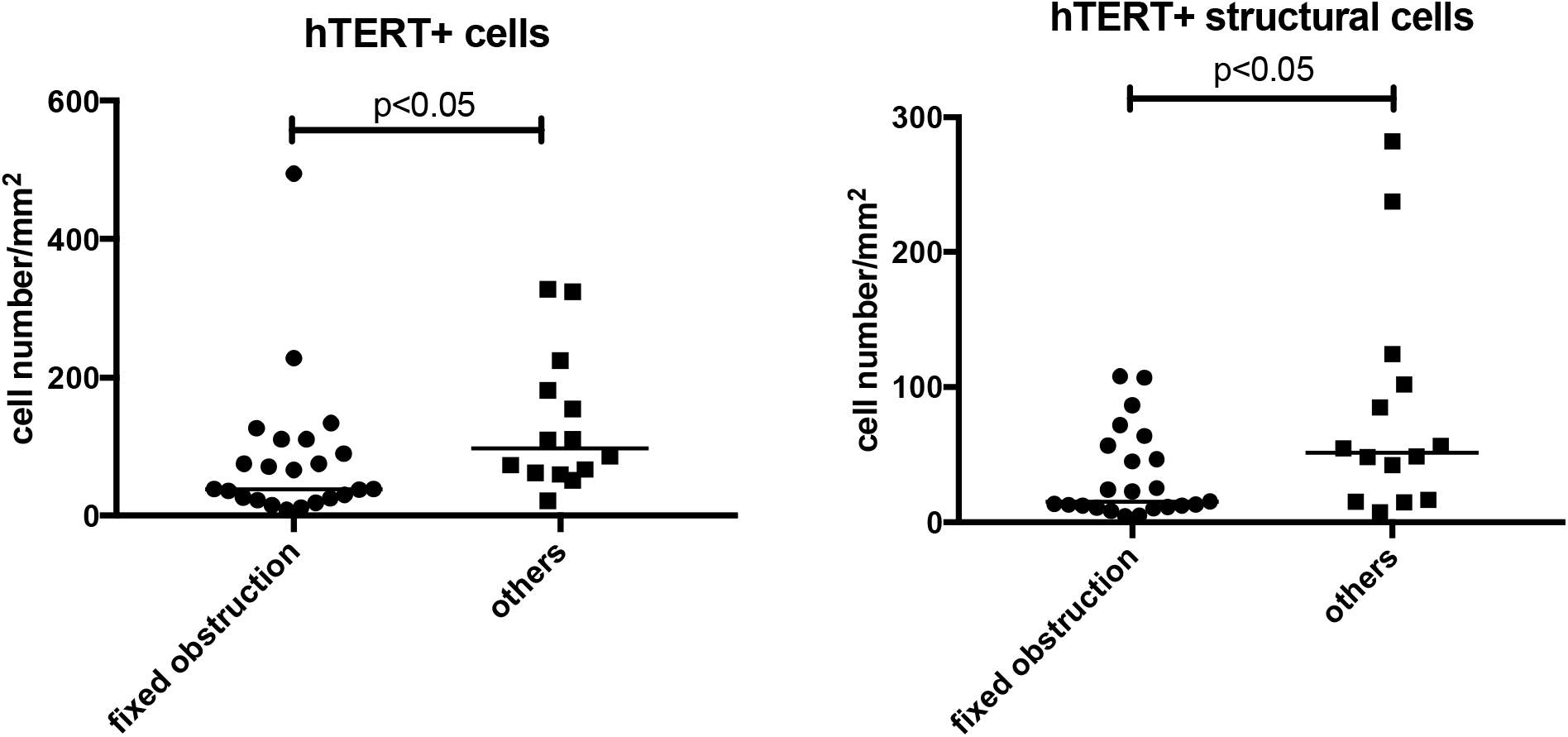
hTERT protein expression in the submucosa of bronchial biopsies of patients with asthma and COPD combined with or without fixed airway limitation. The bar represents the median of the data.

Finally, there was no correlation between hTERT staining for total and immune cells and other lung function parameters such as total lung capacity (TLC), residual volume (RV), carbon monoxide diffusing capacity (DLCO) or carbon monoxide transfer coefficient (KCO).

## DISCUSSION

Our study reports on both global and leukocyte hTERT expression in bronchial mucosa from patients with chronic obstructive airway diseases including asthmatics and COPD and, to the best of our knowledge, is the first to find airway hTERT expression to be inversely correlated with the degree of airway obstruction.

A large meta-analysis found a significant positive association between telomere length in circulating leukocytes and pre-bronchodilation FEV_1_, FVC, and FEV_1_/FVC in asthma and COPD patients [13]. Previous studies have also demonstrated significant telomere attrition in circulating lymphocytes from COPD patients [14,15]. A reduced number of total cells hTERT positive in the submucosa of bronchial biopsies from asthmatic and COPD patients compared to controls has already been shown [8] but there was no assessment of the relationship with lung function.

Our results extend these findings by showing a correlation between hTERT expression in airway mucosa and lung function parameters. We found a positive correlation between airway hTERT expression and FEV1 and FEV1/FVC. Therefore, the more obstructive the patients, the lower the airway hTERT expression. Our finding supports the idea that hTERT expression may protect against fixed airway obstruction in asthma and COPD. However, we did not find any correlation between hTERT and DLCO, which suggests that hTERT in bronchial mucosa may be more involved in airway remodeling than in emphysema, which is associated with severe diffusing capacity impairment. By contrast it was previously demonstrated that alveolar senescence associates with emphysema (13).

Reduced hTERT expression in patients with a fixed airway obstruction was found for the global mucosal expression, mainly accounted for by structural cells which are a combination of fibroblasts and endothelial cells, and a trend was observed for lymphocytes, which is the dominant leukocyte in airway mucosa. Our finding is in keeping with the observation that bronchial fibroblast from asthmatics display telomere shortening as a sign of replicative senescence [16]. Furthermore, by showing that both structural cells and leukocytes from patients with fixed airway obstruction have reduced hTERT expression, we suggest that the lack of hTERT expression may be a general dysregulation in the airways. As telomere shortening was found in circulating leukocytes in other studies with asthma and COPD it further supports the possibility of systemic predisposition as a consequence of environmental and/or genetic factors contributing to telomere erosion and shortening in these patients [17–19]. Why some asthmatics and COPD have altered hTERT expression remains unclear. However, chronic lymphocyte stimulation may play a role. It is well accepted that T-lymphocytes stimulation occurs in the airway of asthmatics and COPD patients as a consequence of chronic inflammation and repeated infections. Lymphocyte stimulation results in loss of telomerase induction which favors telomere attrition [20]. Accelerated senescence of lymphocytes may make the patients with chronic airway diseases more susceptible to infections, which in its turn may aggravate telomere attrition resulting in vicious circle [21–23]. Discrimination of the lymphocyte sub-types that display the lowest hTERT expression was not performed here but warrants further investigation.

Our study has several limitations. First, we did not measure the telomere length. Second, we recruited patients from routine practice who were undergoing endoscopy for other reasons than asthma and COPD. Third, the narrow age range of our cohort made correlation between age and hTERT staining impossible to observe. Fourth, the limited number of patients may have reduced the power of the subgroups analyses precluding the discovery of differences between asthmatics and COPD. Fifth, the patients were receiving maintenance treatment that may have impacted hTERT expression. Indeed it has been shown that ICS may reduce telomere shortening in asthma [24] and the impact of biologics, used in 25% of our asthmatics, are presently unknown.

In conclusion, hTERT bronchial expression in both structural cells and lymphocytes is reduced in severe asthma and COPD who show fixed airway obstruction and could play a crucial role in airway remodeling. Airway hTERT might possibly emerge as a promising therapeutic target in the future to prevent lung function decline in asthma and COPD

## Data Availability

All data produced in the present study are available upon reasonable request to the authors

## Ethic

This study was approved by the Ethics committee of CHU Liege (2015/00c) and all subjects gave written informed consent for participation.

## Author contributions

**CM** designed the study, participated in data analysis and wrote the manuscript; **LM** performed the research and participated in data analysis and manuscript writing; SG participated in data analysis and manuscript reviewing; **NB, FS, JG, J-LC and RL** participated in sample collection and manuscript reviewing. All authors gave final approval of the manuscript and ensured that questions related to the accuracy or integrity of any part of the work were appropriately investigated and resolved.

## Data availability

Data available on request from the authors

