## supplemental material for "Telomerase Reverse Transcriptase Enzyme (hTERT) expression in bronchial mucosa inversely correlates with the severity of airway obstruction in patients with chronic airway obstructive diseases"

Online supplementary material

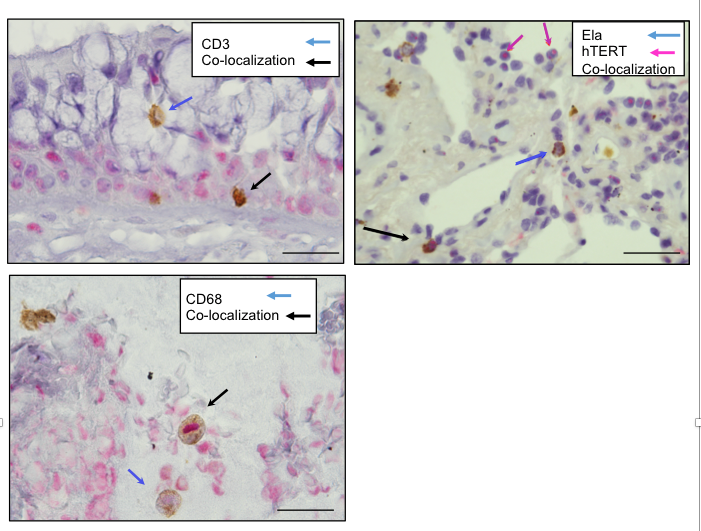

**D**

Eosinophil

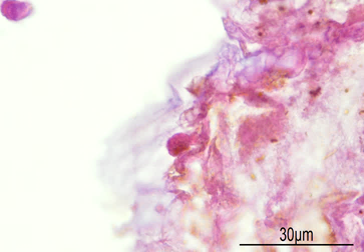

**C**

**B**

**A**

*Figure 1: Immunohistochemistry staining of bronchial mucosa identifying hTERT cellular expression.* ***(A)*** *Co-immunochemistry was made to identify hTERT positive cells and lymphocytes (CD3) (****B****) Co-immunochemistry was made to identify hTERT positive cells and neutrophils (elastase+). (****C****) Co-immunochemistry was performed to identify hTERT positive cells and macrophages (CD68+). (****D****) Immunochemistry was performed to identify hTERT positive cells and eosinophils. The bar represents 30 μm.*
